# Nurses’ and Midwives’ experiences of accessing evidence for teaching, research and clinical practice in Ghana: A mixed methods study

**DOI:** 10.1101/2025.10.15.25337870

**Authors:** Rosie Stenhouse, Joshua Awua, Lawrence Doi, Nancy Innocentia Ebu Enyan, Kingsley Asare, Sarah Amoo, Mengying Zhang

## Abstract

**Aim:** To gain a detailed understanding of N&M experiences of accessing published research to support their practice.

**Design:** Mixed-methods Study

**Background:** Access to up-to-date research evidence is essential for Nurses and Midwives’ (N&Ms) practice, as it enhances patient outcomes. However, N&Ms in Ghana face challenges that limit their ability to access or produce relevant research. It is important to understand the challenges from the perspectives of N&Ms to help in developing effective interventions to enhance evidence-based practice. The six steps to quality intervention development (6SQuID) framework provides a useful framework to engage with N&Ms to identify factors that hinder their ability to access up-to-date research evidence.

**Methods:** A total of 100 participants completed the survey (female 55%) and 43 N&Ms (female, 79%) recruited from two teaching hospitals participated in the workshop to generate the qualitative data. The qualitative data was analysed into themes, quantitative data was analysed using descriptive statistics and integration was achieved through thematic analysis.

**Results:** The results showed that N&Ms face individual level challenges such as insufficient research skills, lack of motivation and have limited time to commit to research related activities. Lack of infrastructure support significantly limit the opportunity for N&Ms access research evidence.

**Conclusion:** The findings point to the complexity of the challenges - occurring across different systems and at different levels - experienced by N&Ms trying to access published research literature in Ghana. Such complexity requires interventions to be developed with a range of stakeholders and focused at different levels from the individual to infrastructural.

**Patient or public contribution:** This study included participatory methods through which the participants defined and framed the problem in a contextually situated manner. However, there was no public or patient involvement in the design, conduct or reporting of this study.

## Introduction

Access to current research evidence is vital for teaching, research, and clinical practice for healthcare practitioners due to the emphasis on evidence-based practice (EBP). EBP is crucial for improving healthcare quality and achieving the United Nations’ (UN) sustainable development goals (UN, 2015) as it is more likely to produce desired patient outcomes in different settings (Connor et al 2023). To maximise the impact of EBP in low-resource settings, nurses and midwives (N&Ms) require access to up-to-date, contextually relevant, and high-quality research evidence (Shayan et al 2019). However, N&Ms from low- and middle-income countries (LMICs) face significant challenges in accessing research evidence because of the limited internet access and institutional resources, high subscription costs for academic journals, and the lack of skills and motivations to search for academic literature (Shayan et al 2019; Alatawi et al 2020; Zhang et al., 2023). One of the possible strategies to address these issues is through strengthening research capacity of N&Ms in LMICs (Zhang et al 2023; Hyder et al 2017; Buser et al 2024).

A concept analysis of research capacity strengthening (RCS) in healthcare settings in LMICs identified three key attributes: individual skill development and training, interdisciplinary collaborations, and the development of contextually relevant research strategies (Buser et al., 2024). Although health challenges are prevalent worldwide, there is a gap in research capacity between high-income countries and LMICs, necessitating more international attention and resource investment in LMICs (McKee et al 2012; ESSENCE 2014; Franzen et al 2017). The need for a contextually-relevant evidence base to enable LMICs to maximise the use of scarce healthcare resources drives the need for RCS in LMICs (Franzen et al 2017; Buser et al 2024). International cooperation is seen as a potential solution to bridge the gap. However, whilst there are guidelines to support good practice in research collaborations with developing countries, such as jointly setting research goals and ensuring both funders and recipients share a voice and ownership of research outcomes (KFPE 2018; ESSENCE and UKCDR 2022), international cooperation is still influenced by power relations (Franzen et al., 2017). In their qualitative meta-narrative review, Franzen et al. (2017) found that whilst there is a shift in power relations toward ‘partnership’, international funders continue to have greater control over the research agenda than local institutions, leading to a lack of sustainable development of research capacity in LMICs.

Buser et al (2024) identify the research-practice gap as an antecedent to RCS, highlighting the need to encourage application of evidence to practice. However, to apply research evidence to practice requires healthcare practitioners to have access to research literature. Our systematic review of the issues experienced by healthcare workers in LMICs when trying to access research literature identified challenges and potential solutions at the individual, institutional, and societal levels (NAMES WITHHELD FOR BLIND REVIEW., 2023). The findings aligned with the ESSENCE (2014) principles of enhancing research capacity, indicating that interventions should comprehensively consider research capacity building at individual, organizational and wider systems levels.

Although this paper presents findings of an empirical mixed methods study to develop a contextual understanding of a sample of Ghanaian nurses’ and midwives’ experiences of accessing research literature, it further provides insight into how these findings have informed the development of a theory of change and further intervention development work with stakeholders aiming to strengthen research capacity of the healthcare workforce in Ghana.

## Methods

The six steps to quality intervention development (6SQuID) (Wight et al 2016) provides a framework for this project (see table 1). This framework takes a co-productive approach recognising that appropriate, acceptable and sustainable interventions are developed through engagement with those people who are experiencing the problem and wider stakeholders who are positioned to influence the factors which are identified as creating or maintaining the problem.

**Table 1:**
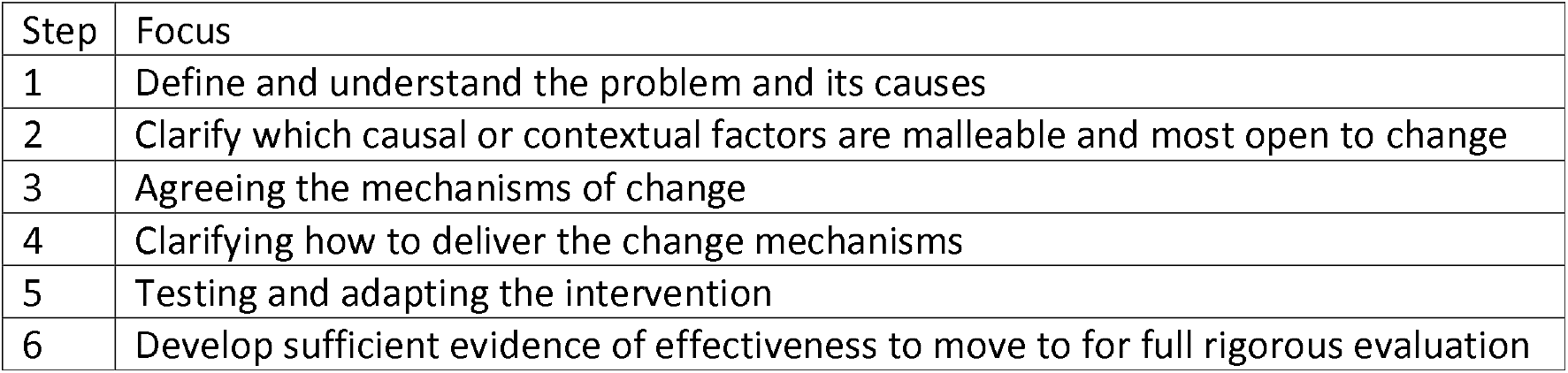
6SQuID steps (Wight et al 2016: 20)

Adopting a co-productive approach is also a way by which we can attempt to decolonise our research relations through attending to relations of power within the research team, in our interactions with potential research sites, and in the design of our data generation activities. The project team is formed of academics from the UK (RS, LD, MZ) and Ghana (NIEE, JA, SA, KA), and the research focus grew out of initial conversations between RS and JA, with ongoing development through discussions across the team as we work. Attentive listening, curiosity, reflexivity and respect underpin our approach.

This project undertook data collection related to the first two steps on the 6SQuID framework leading to a shared understanding of the problem, what creates or maintains the problem, and what may be malleable to change. Data were generated through two workshops and a survey. These two methods were chosen as they provide data in an attempt to understand the breadth of the challenges (using the survey) as well as the gain a deeper understanding of some of the causal mechanisms perceived to underpin these experiences (qualitative workshops). Both forms of data were collected in parallel.

### Setting

Ghana is a West African country which is divided into three zones: south, mid and north. Each of these areas has distinct levels of urbanicity, with the south zone containing the main city of Accra, and some of the coastal towns. The mid-zone is characterised by forestry with its main city of Kumasi. The north has a rural farming area, and unlike the mid and south zones which are majority Christian, it has a large Muslim population. Within Ghana there are 90 health training institutions which provide training to nurses and allied health professionals who graduate with diploma level education. Diploma courses provided by the Health Training Institutions are affiliated to universities and approved by the Director of Health Training Institutions within the Ministry of Health in Ghana. Nursing is offered at degree and masters level by a number of universities in the country. Across both diploma and degree programmes, there are courses on research methods.

The survey was nationally distributed. However, for the workshops, we selected two cities representing the south zone (Teaching Hospital 1) and the north/mid-zone (Teaching Hospital 2) as it was anticipated that the different contexts might create varied accessibility of resources leading to different experiences.

### Participants

Survey participants were recruited through the distribution of the survey link by JA, NIEE, KA through their networks of educators and practitioners. Additionally, a paper version of the survey was distributed to workshop participants.

Recruitment in Teaching Hospital 1 (TH1) was undertaken by the Director of Nursing and SA. In Teaching Hospital 2 (TH2) access was gained through the Director of Research and recruitment undertaken by the Director of Nursing. Each of these gatekeepers had been provided with the participant information sheet to distribute to potential participants. There was enthusiasm for participation in TH1 with a waiting list being formed. To ensure efficient facilitation of the workshops, a total of 45 participants was aimed for across the two sites.

### Data generation

An online survey was developed on the cloud-based platform Qualtrics. Questions were based on the findings of the systematic review (NAME WITHHELD FOR REVIEW 2023) and the experience of Ghanaian members of the research team. Demographic data included the professional discipline of the participant, length of time since qualification, kind of institution in which they worked and zone. Questions focused on where participants accessed research evidence, their literature searching skills and available databases, their use of the internet to access evidence including barriers and common problems, and their knowledge of and engagement with the Health Inter-Network Access to Research Initiative (HINARI) which provides free access to research papers to institutions in LMICs.

Qualitative data were generated through two workshops with nurses and midwives within two teaching hospitals (TH1 and TH2). These workshops were designed to maximise co-production of the definition of the problem, what participants felt caused or maintained the problem and what might be malleable to change. We used small group discussion, fishbone diagrams and other means of generating discussion (see table 2), moving from individual to small group to large group in an attempt to enable those who might be silenced by the power dynamics of the group to have a voice.

**Table 2:**
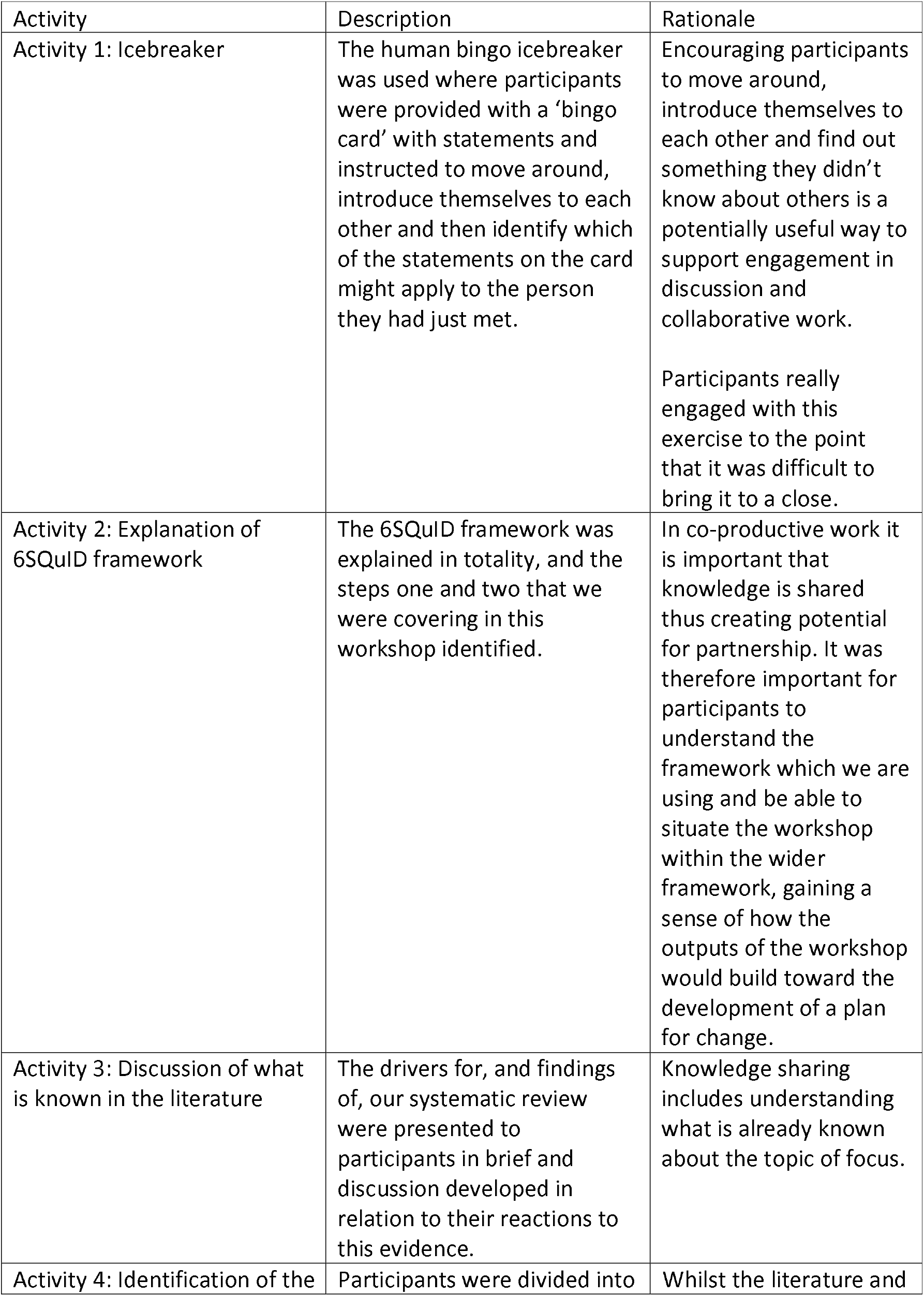
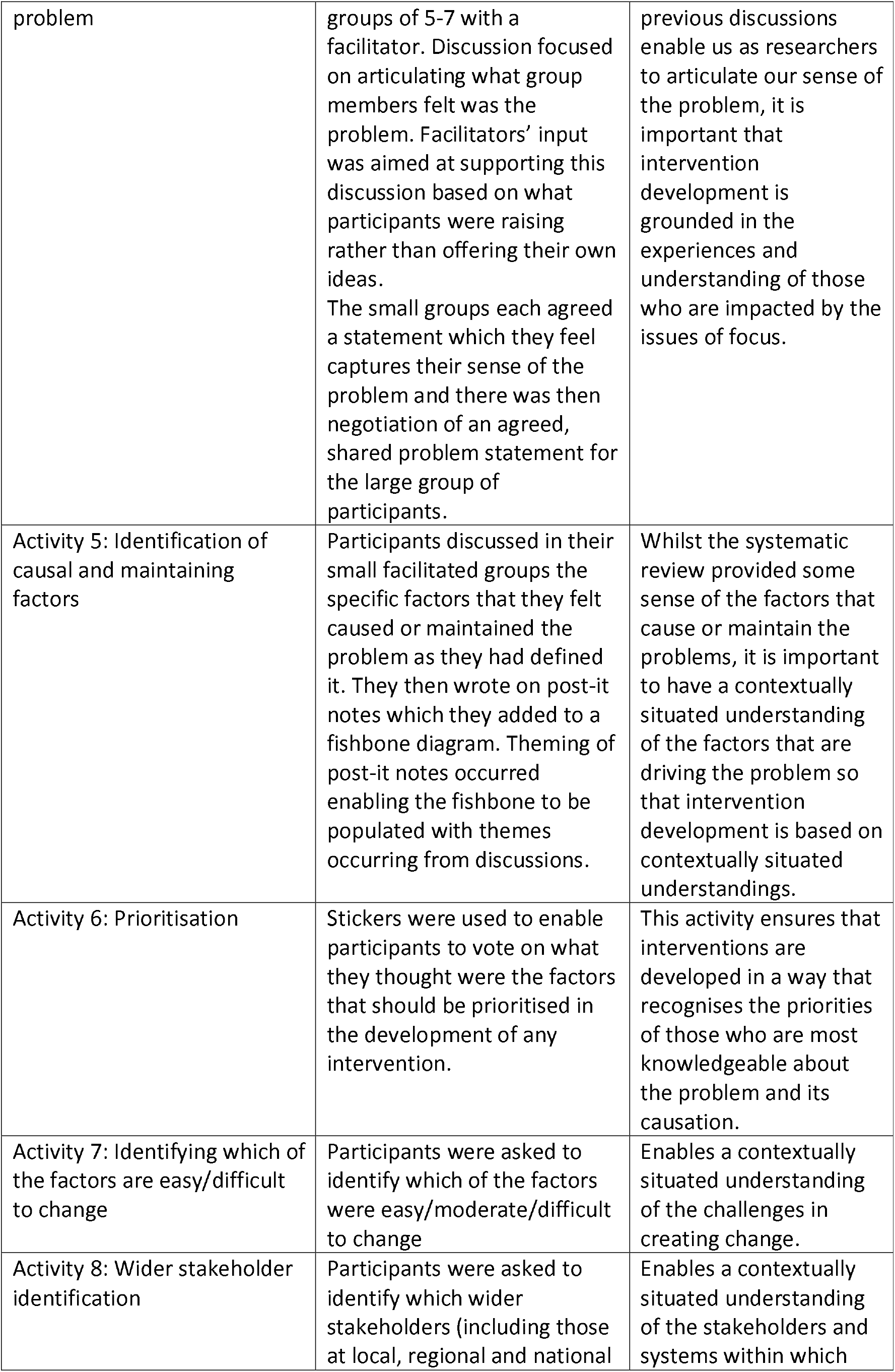
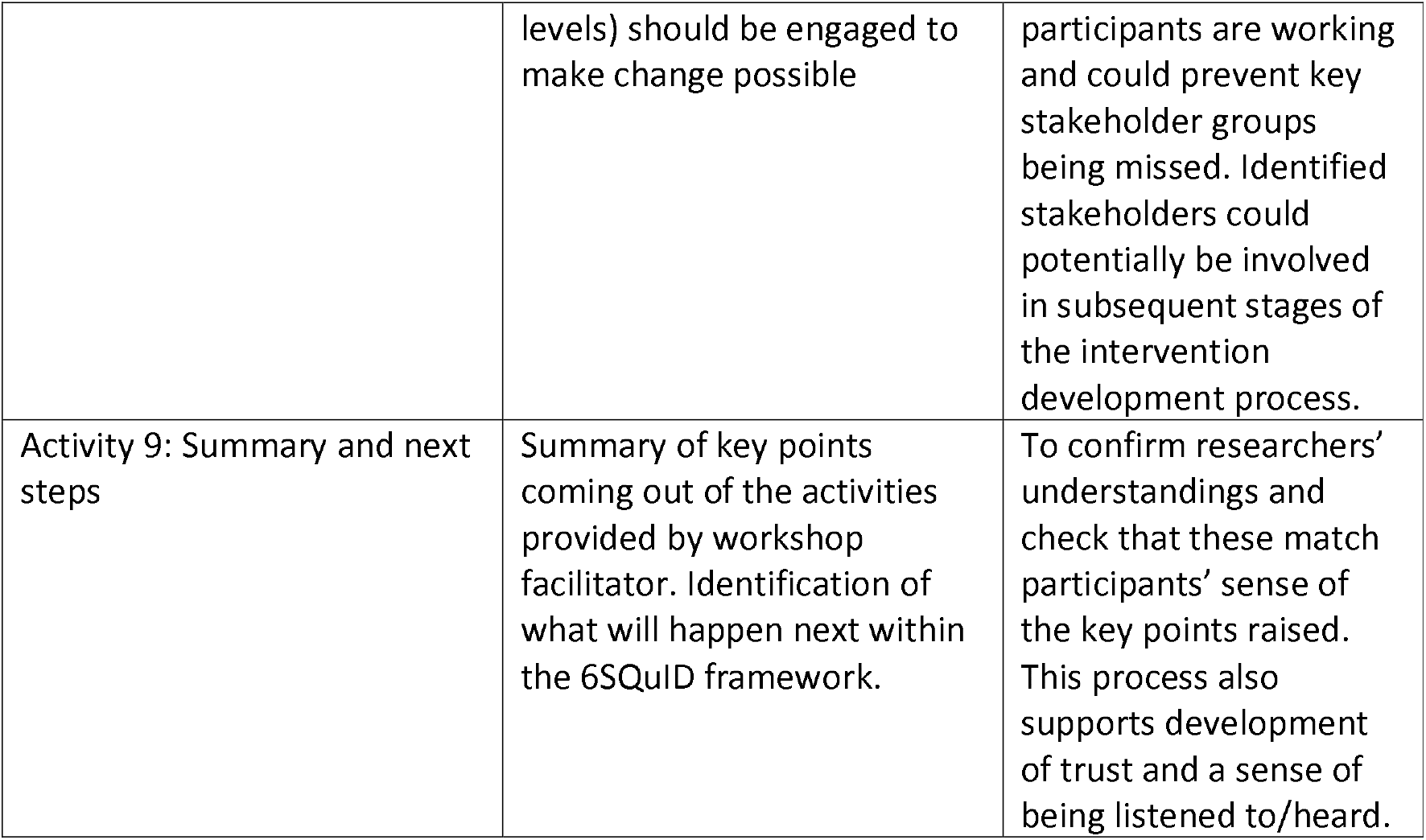
Workshop activity schedule.

**Table 3:** Workshops participant characteristics.

| Site | Male | Female | Years' experience |  |  |
| --- | --- | --- | --- | --- | --- |
|  |  |  | 1-3 years | 4-10 years | 11 years and over |
| TH1 | 5 | 20 | 3 | 15 | 7 |
| TH2 | 4 | 14 | 1 | 7 | 10 |

**Table 4.** Survey participant characteristics.

| Item | N=100 |  |
| --- | --- | --- |
|  | Frequency | Percent % |
| Gender |  |  |
| Male | 45 | 45.0 |
| Female | 55 | 55.0 |
| Age |  |  |
| 21-30 | 25 | 25.0 |
| 31-40 | 57 | 57.0 |
| 41-50 | 18 | 18.0 |
| Profession |  |  |
| General nursing | 63 | 63.0 |
| Midwifery | 12 | 12.0 |
| Mental health nursing | 3 | 3.0 |
| Allied health | 12 | 12.0 |
| Psychology | 2 | 2.0 |
| Other | 8 | 8.0 |
| Workplace |  |  |
| University | 2 | 2.0 |
| College | 15 | 15.0 |
| Hospital/clinic | 83 | 83.0 |

Data took the form of collective statements and points made during the activities 4 to 8 (table 2). Fieldnotes, in the form of reflections on the process and discussions, were made by workshop facilitators (RS, JA, LD, NIEE, MZ, SA, KA).

### Data analysis

Workshop data were transcribed from the workshop materials such as post-it notes, flip charts and fishbone diagrams, into a word document for each of the workshops, and these were brought together to provide an overall sense of convergence and divergence of experiences and opinions in the two workshop settings. Descriptive statistics were conducted using SPSS. Integration of the qualitative and quantitative findings was carried out through a process of thematic development by LD and RS, confirmed by JA, NIEE, MZ. The thematic analysis was guided by the socio-ecological model of health (Dahlgren & Whitehead 2021) and conceptualised as individual, institutional and infrastructural/societal factors. As with Dahlgren and Whitehead (2021), connections between the different levels were made clear by the participants in some of the workshop discussion and these perceived connections were captured in the fieldnotes and guided the analysis.

### Ethics

Ethical approval was granted by the School of Health in Social Science, University of Edinburgh in the UK and by the two teaching hospitals in Ghana. The Ministry of Health, Ghana also gave administrative approval. Participation was voluntary and participants provided written informed consent to participation in the workshop. Participant information and consent were included in the survey, with a statement at the end of the survey reminding participants that submission of their completed survey indicated consent to participate. Data were managed in line with University of Edinburgh data management and data protection policies. Survey data were anonymous and aggregated, and workshop data were the product of group discussion and therefore co-constructed and unattributed, supporting participant anonymity. Given the potential sensitivity of the study focus, the settings are also anonymised.

### Findings

#### Participant characteristics

Participants at the workshop at TH1 were 25 nurses and midwives from a range of clinical areas.

Participants in the workshop at TH2 were 18 nurses from a range of clinical departments including three in senior nurse management positions.

A total of 100 health professionals completed the survey questionnaire. The profile of the professionals who took part in the survey is shown in Figure 1. More females (55%) than males (45%) responded to the survey. This is expected given that the nursing and midwifery profession in Ghana is female dominated. Of the total respondents, 63% were general nurses and only 2% were psychologists. Majority of the respondents were hospital or clinic workers (83%).

**Figure 1:**
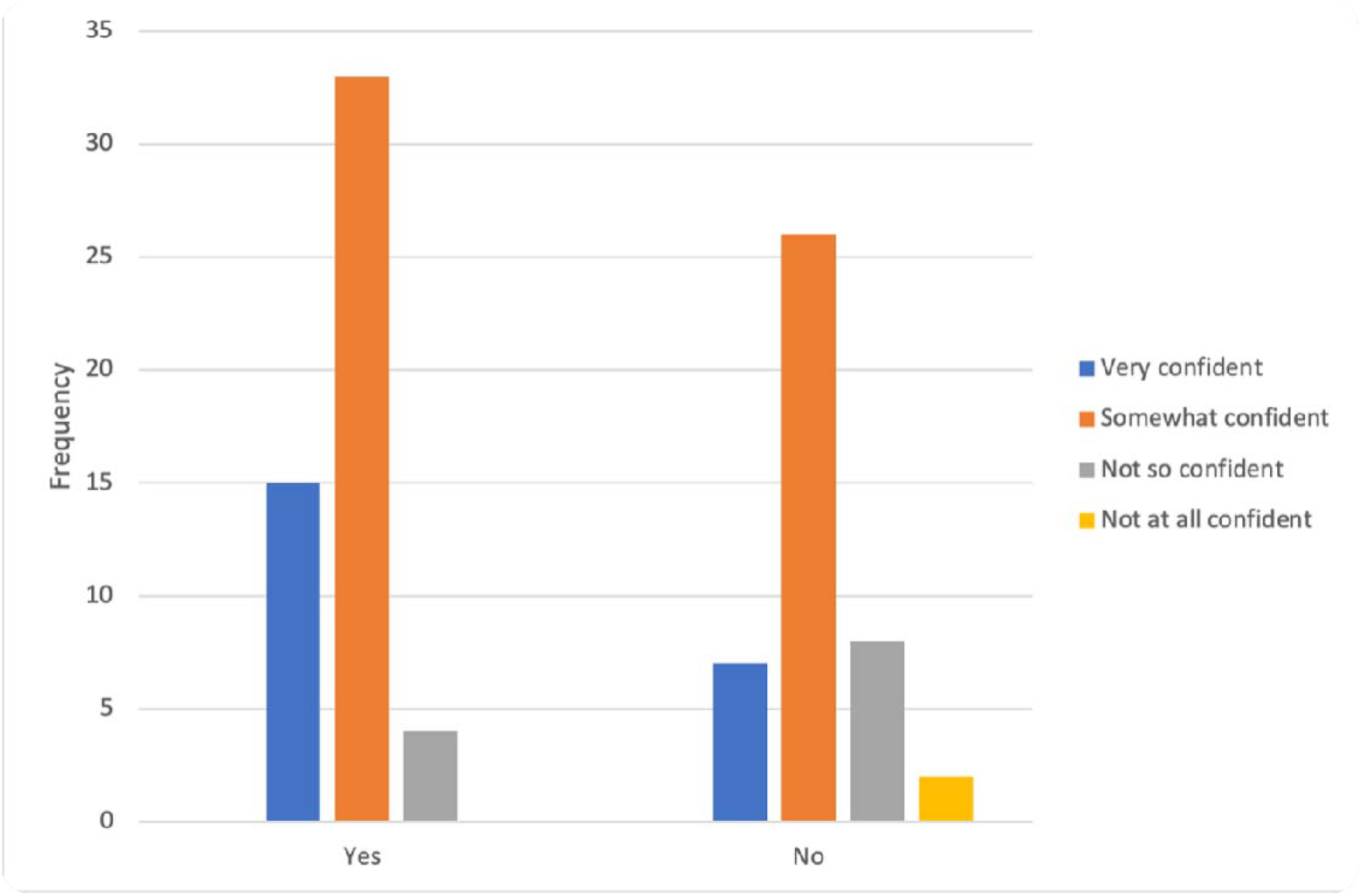
Confidence in searching for literature online between those who have had formal training and those with no formal training.

#### 6SQuID step 1: Identification of the problem

Workshop participants were asked to identify and form a consensus around what they felt was the main problem that needed tackling. Whilst there were some differences between workshops, the combined information from the workshops identified a multifaceted problem involving:

- Insufficient practical training in research including literature searching skills
- Insufficient interest in research
- Insufficient institutional and infrastructural support for research including network issues, affordability of resources

#### 6SQuID step 2: Causal factors and factors maintaining the problem

The groups identified a range of causal and maintaining factors across the two workshops. This data is presented alongside the data from the survey in line with a parallel mixed-methods design. Factors are presented as individual level factors; institutional level factors; and infrastructural level factors.

##### Individual level factors

Insufficient research skills or competence: Workshop participants perceived that this was generally associated with “*lack of practical research training*” including literature searching and reviewing, and “*inadequate opportunity to engage in research trainings* [sic]” or capacity development activities through engagement with research teams in the clinical areas.

In the survey, when respondents were asked to indicate whether they have had any formal training on searching for literature online, 56% (n=55) responded in the affirmative, with 44% responding no (n=99; data missing n=1). When those who responded in the affirmative were invited to rate the formal training they had received, 61% said the training was very or quite comprehensive and 39% felt it was not so comprehensive (n=52; data missing n=3).

When respondents were asked to indicate how confident they were with searching for literature online, 23% said they were very confident, 62% were somewhat confident, 13% were not so confident and only 2% were not at all confident (n=95; data missing n=5). When comparing those who have had formal training with those who said they have not had any such training, it was clear that those who have had formal training were more confident than those who have not had training (see figure 1).

###### Lack of personal motivation

Workshop participants expressed that whilst they were motivated to engage with research literature and EBP, they felt that others in the organisation did not value or were not motivated to engage with EBP because of their *“poor orientation and mindset toward research”*.

###### Lack of time

Literature searching was understood to be time consuming and there was limited time, aside from clinical practice that could be spent doing this.

##### Institutional level factors

Insufficient institutional value placed on EBP and nursing research: Workshop participants were enthusiastic about engagement with research. However, they sensed that their institutions did not place value on locally developed research evidence as a means of shaping clinical practice and that this demotivated staff: *“non-recognition of the impact of research findings in the facility. No practical application of* [local] *research findings”*.

Insufficient institutional access to research literature: Workshop participants identified that their institutions provided inadequate access to up-to-date full text journals in the library. HINARI provides free access to online health research literature and is available to health professionals and academics working in institutions in LMICs (Robertson 2014; <u>Hinari -</u> Research4Life). When participants were asked to indicate whether they have heard about HINARI, 37% (n = 38) responded ‘yes’ and 63% said ‘no’ (n=64). Amongst those who had heard of HINARI, we asked them to indicate whether they had used HINARI to access health literature before or not. Sixty-five percent had used HINARI, with 27% using it in the past three months. Respondents also responded to a question about how easy it was to obtain login details or password from their institution to access HINARI. Majority (62%) felt it was extremely or somewhat difficult, with 33% responding that it was extremely or somewhat easy as shown in Table 5.

**Table 5:** Experiences of using HINARI.

| Item | Options | Frequency | Percent (%) |
| --- | --- | --- | --- |
| Used HINARI to access health literature? (n=34) | Yes | 22 | 65 |
|  | No | 12 | 35 |
| How often have you used HINARI to access health literature? (n=22) | <3months | 6 | 27 |
|  | >3months | 15 | 68 |
|  | Never | 1 | 5 |
| How easy was it to get login/password details from your institution? (n=21) | Extremely or somewhat difficult | 13 | 62 |
|  | Neither easy nor difficult | 1 | 5 |
|  | Extremely or somewhat easy | 7 | 33 |

###### Research skills development during first level N&M education

Workshop participants identified how the lack of research activity of the staff who teach research methods to N&Ms during their pre-registration programmes led to this teaching being more theoretical than practical: *“those teaching research in institutions are not researchers themselves so struggle to explain the concepts/skills etc”*. They felt that research skills teaching should be *“practical based, not only theoretical”*. They felt this lack of practical engagement with research demotivated students in relation to engaging with research, and that this was connected with the lack of research engagement that they experienced from colleagues. Participants felt that generally N&M pre-registration education does not provide the skills to enable qualified staff to conduct research.

##### Infrastructural/societal level factors

###### Poor internet access

Many participants at the workshops cited poor internet access as a problem in terms of quality and access permissions in clinical areas: *“fluctuations and poor quality internet coupled with restrictions on access to certain websites”*.

In the survey, it was clear that most respondents used their personal data to access health literature. When asked that on a typical day when accessing or downloading literature from the internet, what internet sources did they use, 93% stated that they used personal wifi or data from their phone or tablet. Only 7% used wifi from their institution as depicted in Figure 2 (n=96).

**Figure 2:**
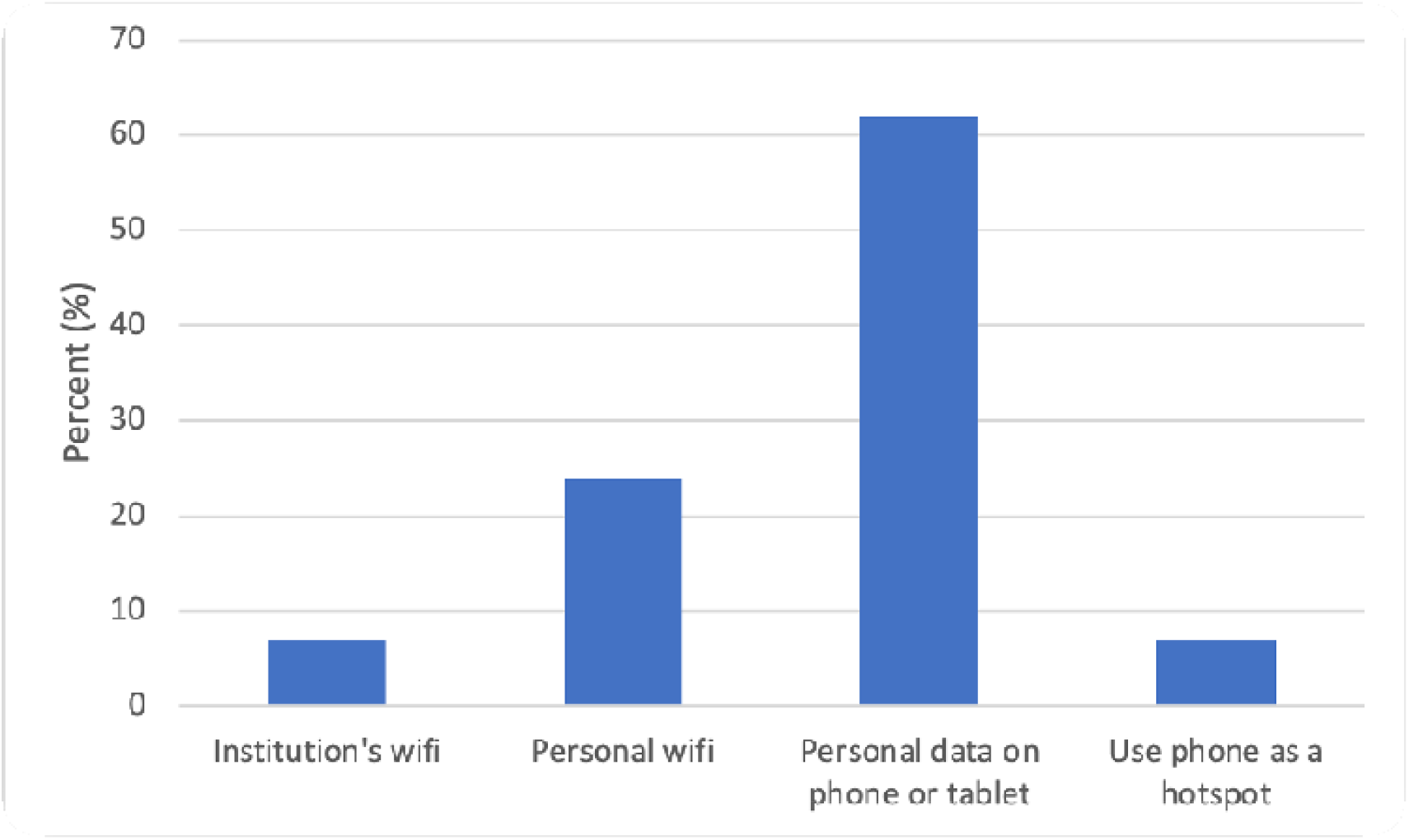
sources of internet to access literature.

When respondents were asked to identify the key barriers to accessing literature online, cost of mobile data (54%), website requiring payment (48%), poor wifi (34%), no access to wifi (26%) and inadequate research skills (23%) were identified as barriers (respondents could select up to four barriers).

## Discussion

This study used a mixed-method approach to further our understanding of N&Ms’ experiences of accessing published research in Ghana. The findings showed that N&Ms face challenges at multiple levels; individual, institutional and infrastructural affecting their ability to access published research. These findings can be understood in the context of research capacity strengthening with access to published research forming a foundation block upon which to build.

On an individual level, the results from the study showed that N&Ms have insufficient research skills, lack motivation and have limited time to commit to research related activities. These findings are consistent with previous studies that reported lack of skills and time for research as significant barriers to N&M EBP and research production (Cheng et al., 2017; Ogundahunsi et al., 2015; Trotter and Kell, 2014; Zhang et al., 2023). The survey findings indicate that search skills training may increase confidence which is in line with the findings of Garg et al’s (2003) critical appraisal of the literature which found only a weak positive impact of one to eight hours of search and retrieval skills training. However, Hatahet et al’s (2023) novel approach using peer group mentoring between post-graduate students in a high-income university and undergraduate students in Syria found a significant impact on skills development. This may indicate that the pedagogy of literature search skills training should be considered at the design stage.

Research skills such as literature search skills are also part of the N&M pre-registration educational curriculum in Ghana with all students undertaking a research course and a research project prior to graduation. The findings that student nurses are demotivated in relation to research skill development is a recognised challenge for nurse educators (Niven et al 2013) as students value their clinical skills over their research skills. However, more recent evidence identifies that undergraduate nursing students in two universities in the UK and Slovenia recognised the importance of EBP and research for their clinical practice (Brooke et al 2015). Our finding that there was a perceived lack of motivation within qualified staff groups to engage with research literature indicates how such student disengagement might carry through to the qualified workforce, a finding supported by Ryan’s (2016) integrative literature review. Given the high patient to N&Ms ratio in Ghana (1 nurse to 839 population in Ghana in comparison to 6.66 qualified nurses per 1000 population in the UK) (GhanaWeb 2019; Nuffield Trust 2024), it is reasonable that N&Ms prioritise patient care over searching for or engaging in/with research. However, there remains a professional imperative, perhaps more so in low resource settings (Buser et al 2024), to access and generate contextually appropriate evidence (Boaz et al 2024). Creating cultures in LMIC healthcare institutions which value N&M research is central to RCS (Buser 2024). Increased visibility of nursing and midwifery research within the educational and clinical contexts can help create such cultures (Scala et al 2016) which can increase student and qualified N&Ms’ understanding of, and engagement with, the role of research as a normative part of N&M work (Henshall et al 2024; Scala et al 2016). This points to the importance of RCS work where nurses are visibly leading and undertaking research in clinical areas.

Many institutions in LMICs have difficulty accessing research evidence partly due to limited financial resources, and lack of access to databases (Annan, 2004; Paci et al., 2021; Singh et al., 2011; Young et al., 2016; Zhang et al., 2023). Whilst increased confidence and competence in literature searching skills might increase the ability to find citations and abstracts of published literature, the difficulties accessing full-text papers is indicative of a need to increase availability of these online. HINARI offers a freely available opportunity to increase the accessibility of full-text resources in LMICs such as Ghana (Robertson 2014). Despite this, only 47 of the 92 Health Training Institutions in Ghana are registered with HINARI (<u>Research4Life Registered Academic Institutions</u>). Increasing search skills without improving access to sources of full-text literature that is not behind a paywall will not increase access to published research, and may lead to demotivation if searches lead to inaccessible literature.

Infrastructural challenges such as access to the internet also interfered with the use of research in nursing practice. Within our data, over 90% of the respondents relied on their own internet to access online materials due to the lack of institutionally available wifi. Internet data is costly and not everyone can afford. Infrastructural level challenges have been reported in prior studies (Smith et al., 2007; Young et al., 2016) and add to the complexity of the challenges experienced by N&Ms trying to access published literature in LMICs.

### How the findings informed a logic model development

Intervention development involves theorising how the most modifiable causal factors could be changed to lead to intended outcomes (Wight et al., 2016). The use of programme theory, for instance in the form of a logic model to articulate how change can be affected can promote shared understanding of the intervention amongst stakeholders (Skivington et al., 2021). Following developing a contextual understanding of the experiences of accessing research literature and in line with the 6SQuID framework, we invited stakeholders including N&Ms and decision makers at the ministry of health to take part in developing a programme theory (step 3 of 6SQuID). To develop the logic model, the research team undertook a preliminary literature review to understand the evidence underpinning how change could come about for each of the modifiable factors identified in step two of 6SQuID. This led to the development of an initial logic model. This was presented to the stakeholders in an online workshop. Stakeholders discussed and mapped out the activities that could trigger change at the short, medium and longer-term to address the identified issues associated with inability to access contextually appropriate literature to inform N&M practice (Figure 3). The next step of the project will involve working with stakeholders to identify activities and actions to deliver change or ‘activate’ the programme theory. This will lead to the development of an implementation plan (step 4 of 6SQuID).

**Figure 3:**
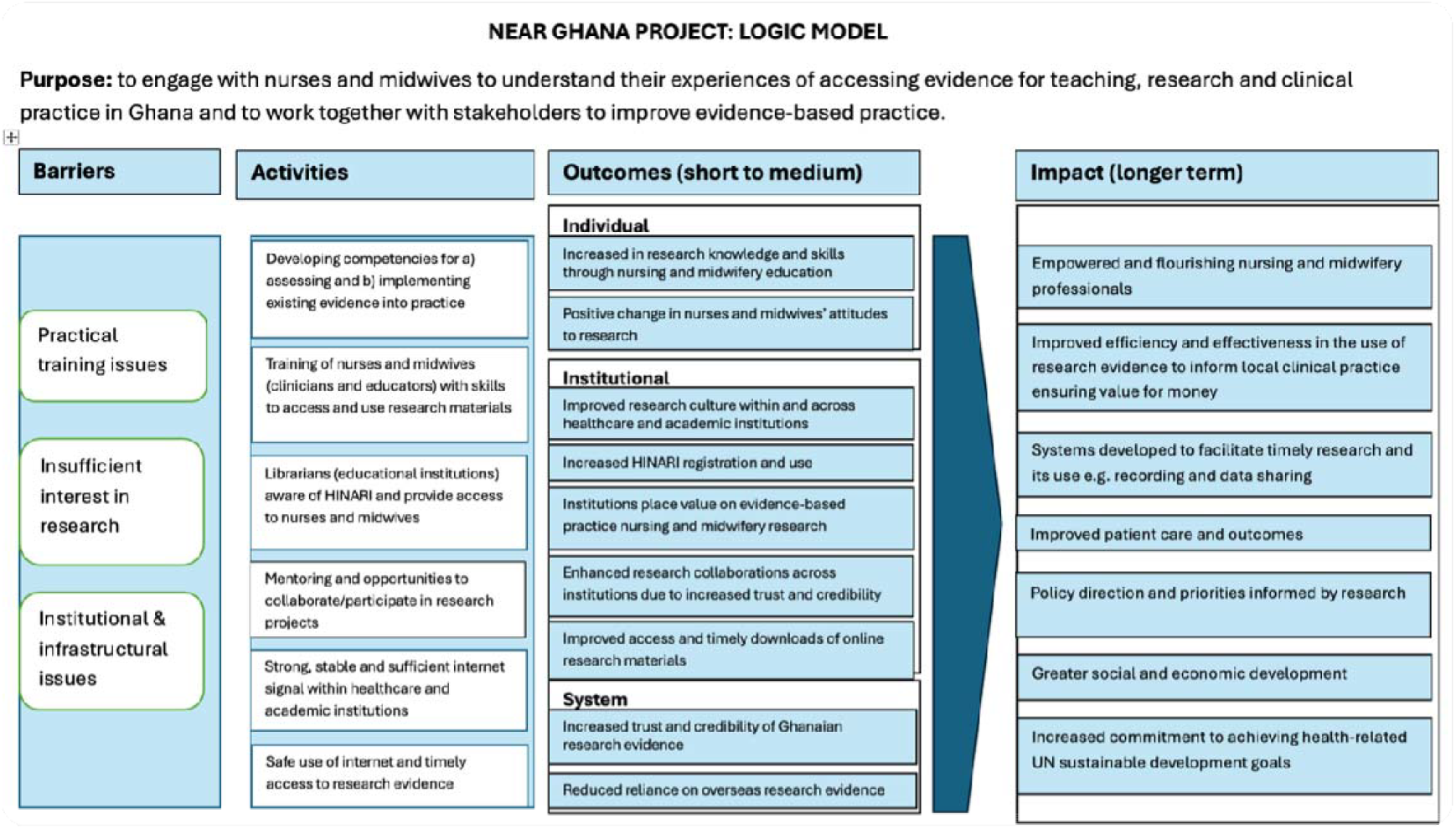
Logic Model of NEAR Ghana project.

## Strengths and Limitations

This study provides an insight into the challenges experienced by a small geographically limited sample of N&Ms in Ghana. The participants who elected to join the workshops were highly motivated in relation to research and may well not represent the wider N&M workforce. However, the findings from both quantitative and qualitative data built a coherent picture, providing a stronger evidence base than each dataset on its own. On the basis of the small sample sizes, the findings are not generalisable, but may be transferable to other contexts. However, these findings add to other research within LMICs in Africa building a body of evidence. Further research across Ghana and other sub-Saharan African countries is required to understand the situation across a wider geographical area as well as identify strategies that have been effectively implemented in these settings to strengthen research capacity. This study focused only on understanding issues involved in accessing published research literature to support N&Ms practice and identified factors that contribute to these issues. This is an ongoing research project, and the research team are currently working with stakeholders in Ghana to identify priorities for change and ways of delivering change to enhance N&Ms’ EBP and research capacity.

## Conclusions

The findings point to the complexity of the challenges experienced by N&Ms trying to access published research literature. Such complexity, where challenges occur across a range of systems and at different levels, requires interventions focused at different levels from the individual to infrastructural. Such interventions require engagement with a range of key stakeholders who are in a position to support and maintain sustainable change. The recent Nursing and Midwifery Strategic Plan (2024-2028) published by the Ministry of Health in Ghana has a clear focus on research capacity strengthening creating the opportunity for intervention development and implementation.

## Data Availability

All data produced in the present study are available upon reasonable request to the authors

